# #ActuallyAutistic Twitter dataset for precision diagnosis of Autism Spectrum Disorder (ASD)

**DOI:** 10.1101/2023.09.19.23295799

**Authors:** Aditi Jaiswal, Peter Washington

**Affiliations:** Department of Information and Computer Sciences, University of Hawaii at Manoa

**Keywords:** Social media analysis, Twitter, autism spectrum disorder, sentiment analysis, public health

## Abstract

The increasing usage of social media platforms has given rise to an unprecedented surge in user-generated content with millions of users sharing their thoughts, experiences, and health-related information. Because of this social media has turned out to be a useful means to study and understand public health. Twitter is one such platform that has proven to be a valuable source of such information for both public and health officials. We present a novel dataset consisting of 6,515,470 tweets collected from users self identifying with autism using “#ActuallyAutistic” and a control group. The dataset also has supporting information such as posting dates, follower count, geographical location, and interaction metrics. We illustrate the utility of the dataset through common Natural Language Processing (NLP) applications such as sentiment analysis, tweet and user classification, and topic modeling. The textual differences in social media communications can help researchers and clinicians to conduct symptomatology studies, in natural settings, by establishing effective biomarkers to distinguish an autistic individual from their typical peers. For better accessibility, reusability and new research insights, we have released the dataset publicly.

## Background and Summary

Autism spectrum disorder (ASD) is a developmental delay causing physical, cognitive, and behavioral changes and affecting millions of individuals. A core complexity of ASD lies in its symptom profile changing with age, often leading to the misattribution of its characteristics to other conditions such as anxiety, obsessive-compulsive disorder (OCD), and attention-deficit/hyperactivity disorder (ADHD)^1,2^. Therefore, an early diagnosis is crucial to provide appropriate treatment and improve the efficacy of screening tools. However, there are limitations on the availability of standard tests^3^, leading to misdiagnosis or delayed treatments^4^, which can place patients at risk of developing depression or suicidal tendencies^5^. Social media has turned out to be a useful means for real-time public health monitoring. Such non-clinical data holds considerable potential for the research community to extract meaningful insights through a less intrusive approach and improve the rigor of ASD analytics research. The digital footprint of an individual can be analyzed to study behavioral symptoms of ASD and other mental health disorders^6^.

Twitter, a popular microblogging platform, has emerged as a particularly valuable source for data. The platform allows users to post tweets containing up to 280 characters and has an active monthly user base of approximately 450 million individuals^7^. Twitter’s strength lies in its ability to capture real-time thoughts, news, conversations, and statistics, making it more suitable for collecting observational data than traditional survey-based methods. Research in mental health such as identifying depression and mood changes^8–14^, and real-time mapping of natural disasters^15,16^ or infectious disease spread and its effect on emotional health^17–24^ has greatly benefited from digital phenotyping.

ASD has been the subject of multiple clinical trials, reviews, and epidemiological studies conducted using behavioral features such as eye gaze^25^, prosody^26^, asynchronous body movement^27^, facial expressions ^28,29^, mobile phone data^30–33^ or even electroencephalogram (EEG)^34^. However, only a handful of studies have used social analytical tools^35–38^, especially using Twitter^39,40,41^ for investigating ASD. In addition, other social networking sites such as Reddit^42–45^, Facebook^46^, Instagram^47,48^, Flickr^49^ and Sina Weibo^50^ have also provided a valuable source of data for detecting and studying mental health conditions, substance abuse and risky behaviors. Using these prior works as inspiration, we curated a novel large scale Twitter dataset to study various aspects of social communication that differentiate autistic people from their neurotypical peers. Figure 1 provides an overview of the steps taken to curate the dataset.

**Figure 1.**
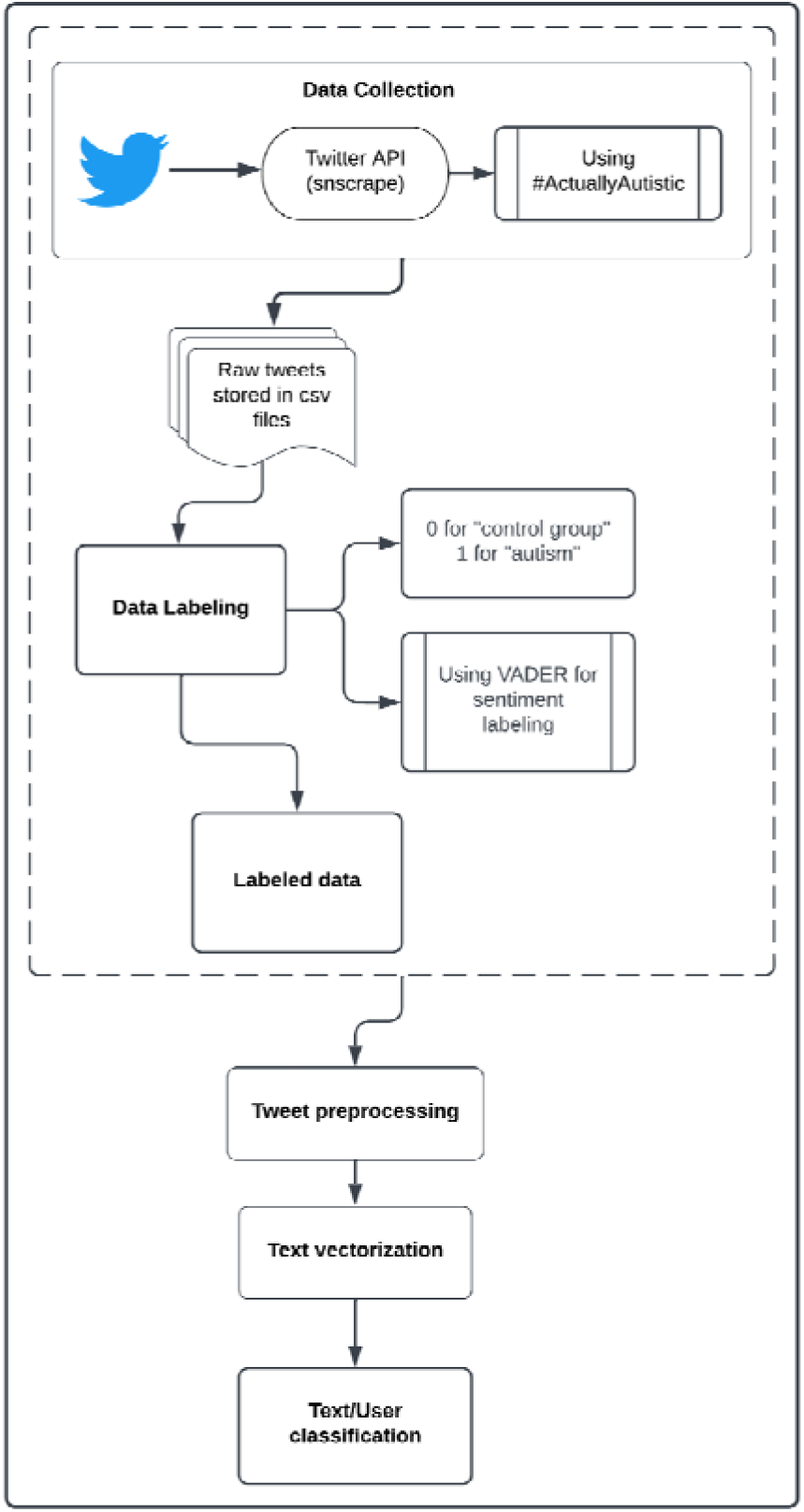
Pipeline for creation of the novel Twitter autism dataset.

The profound shift in society’s reliance on social media for information, in contrast to traditional news sources, coupled with the enormity of data generated, has brought about an increased focus on the use of NLP for text analytics. Although research tools using facial expressions^6,51–60^ and eye gazing for phenotyping ASD^61,62^ are consistently reliable, there is currently a lack of standardization and preciseness in tools to measure deficits in social interaction. Therefore, linguistic and behavioral markers extracted from Twitter conversations can be used to study textual differences and social interactions in naturalistic settings. This dataset, which we release to the public, can be used by researchers and clinicians to understand and analyze textual different features associated with ASD, enabling the research community to build precision health tools to identify and monitor ASD and its early symptoms, identify specific behavioral characteristics, derive hidden patterns, propose a clinical treatment plan, and provide community support. Furthermore, sharing the data can promote interdisciplinary collaboration to gain fresh perspectives on the research problem, and promote awareness and solution finding through hackathons, tutorials, and public challenges.

## Methods

### Data Collection

In recent years, hashtags such as #MeToo, #BlackLivesMatter, and #StopAsianHate has played a significant role in promoting social movements and campaigns, including those aimed at raising awareness about specific societal issues. Within the ASD community, the popular hashtags were #AutismMom and #AutismParent, representing neurotypical parents of autistic children whose outside perspectives have often shaped research and policy in this area. However, these advocacy groups often overshadow autistic adults, creating a gap in the decision-making process. To address this issue, a paradigm shift occurred in the autism rights movement through the hashtag ‘*#ActuallyAutistic*’, focusing on understanding the experiences, challenges, and lives of individuals on the autism spectrum rather than solely focusing on caregivers. We extracted Twitter conversations of users self-identifying with ASD using this hashtag to study the differences in linguistic patterns of autistic people.

We collected tweets using *snscrape*^63^, a Python based library allowing social media scraping without the need for personal Twitter API keys which provides a powerful search functionality to help filter tweets based on various conditions, such as date-time, language or location. We specifically targeted English tweets using the search query ‘*#ActuallyAutistic*’ ranging from January 1, 2014 to December 31, 2022. From these tweets, we identified unique users who had keywords like “autism” OR “autistic” OR “neurodiverse” in their profile description (i.e., Twitter bio), signifying self-identification with ASD. It is worth noting that some users only had these keywords in their username, so we also examined usernames and their tweet contents in addition to user bios. Finally, we extracted all the tweets from the timelines of these users to construct the autism dataset, which consists of 3,137,952 tweets from 17,323 individuals. Associated metadata such as username, account created, friends count, date of tweets posted, and location (if the user had mentioned in their profile) were also extracted that could be used for statistical or network analysis.

To build a tweet classifier between individuals with ASD and their neurotypical peers, we collected a sample of random tweets as a part of a control group. To achieve this, we formulated a search query excluding the hashtag i.e. “*-#ActuallyAutistic*”, using the advanced query searching operators and methods provided by Dr. Igor Brigadir^64^. However, this approach carries the risk of data leakage, whereby users who have not posted any autism-related content may possess autism-related keywords in their profile description or username. To avoid this, we screened users who had any such keywords in their profile description or usernames, or who were also present in the autism dataset, and subsequently removed them from the sample. As the main objective of curating a control group dataset was to have tweets different from those posted by ASD users, and given that there are millions of tweets posted everyday, we collected 1,000 tweets per day between the same time period to build the control group dataset. Through this, we obtained 3,377,518 tweets from 171,273 individual users.

### Data Labeling

To effectively train a supervised machine learning model, it is necessary to have labeled data, where each data point is associated with a corresponding class. We manually annotated the tweets posted by individuals with ASD as belonging to the class “autism”, assigned label 1. All other tweets were labeled as belonging to the class “control group” or label 0. It is important to note that obtaining ground-truth labels can be a costly and time-consuming process, and performance of the machine learning model is often found to decrease with a decrease in labeled data. Weak supervision approaches leverage partially accurate or noisy sources for annotations, which can be more efficient than manual labeling.

Below is a small sample of tweets in our dataset and their corresponding labels:

• Sadly as #autistic #adhd #audhd we have limited choices in life and interm of profession and other life circumstances. I wish we have friendly #Neurodivergent environment in work place, where we have a sensory room too for #ActuallyAutistic #actuallyadhd . (Labeled as belonging to class “autism”)
• Celtic are now only 3 trophies behind the ‘World’s most successful team’. And they are 9 pts ahead for another Let that sink in This time next year they could be level on trophies won History being destroyed by this board; players (Labeled as belonging to class “random”)

This study has been approved by the University of Hawaii Institutional Review Board (UH IRB) under an expedited review procedure and the user information has been deidentified.

### Data Preprocessing

Working with raw, unstructured Twitter data is challenging because the conversational text has too many noisy elements such as punctuation, abbreviations, emojis and other stray characters. Thus, before using such data for model training, it is necessary to clean and preprocess the data, which is an essential step for any NLP task. We started by removing the usage of any profane language in the tweets such as cursing or swear words using a Python library called *better- profanity*^65^, which is designed to flag inappropriate words using string comparison and mask them using special characters (the default setting uses “*”). We then tokenized the text into words, removed any non-alphanumeric characters, hyperlinks, user mentions, and HTML tags, and converted the word tokens into lower case to avoid any confusion and data redundancy. We then removed stop words to avoid adding noise and complexity to the features with no meaningful information. To further simplify the input space and normalize the vocabulary, we applied stemming and lemmatization. We also removed any hashtags or a list of keywords related to ASD such as ‘actuallyautistic’, ‘autism’, ‘autistic’, ‘autismacceptance’, ‘autismawareness’, ‘askingautistics’, ‘askingautistic’, ‘neurodiversity’, ‘neurodivergent’, ‘allautistics’, ‘adhd’, ‘mentalhealth’, ‘asd’, ‘diagnosis’, ‘autistics’, ‘autismpride’, ‘autismspeaks’, which could introduce bias and lead to model overfitting.

### Text classification

To build a tweet classifier, we first identified unique users from both the ASD and control datasets and split them into a 85:15 ratio for training and testing. This was done to avoid data leakage, which could occur if any user’s tweets were split between the training and testing sets, causing the model to overfit by learning the semantic patterns specific to an individual user. The tweets were preprocessed as defined in the previous section and formed the training and test dataset. The categorical labels, representing whether a tweet belonged to an ASD or control user, were used for model training and evaluation. The training dataset was further divided into 85% training data and 15% validation data, which was used to fine-tune the model and adjust hyperparameters.

For text to numeric vectorization, we used term frequency-inverse document frequency (TF-IDF) as well as a predefined word2vec embedding method. We started by training TF-IDF word vectors using various classical ML algorithms: Support Vector Machines (SVM), Naive Bayes (NB), Logistic Regression (LR), and XGBoost gradient boosting (XGB), using 5-fold cross-validation and accuracy as the primary evaluation metric to identify the best classification method. We then trained the word2vec model using the best identified algorithm for better feature representation capturing the semantic and syntactic similarity of words and measured a more comprehensive list of evaluation metrics.

### User profile classification

To further evaluate the effectiveness of the curated dataset, we performed user classification using all the tweets from each user’s timeline. We started by identifying all the unique users who had posted at least 5 tweets and divided them into an 80:20 training to test ratio in order to avoid model overfitting as users with only one tweet might not be representative of the population. The preprocessed tweets from each user were then grouped together to form an individual document. For model training, we used an attention-based bidirectional long-short term memory (Bi-LSTM) model vectorized with a randomly initialized, self-trained embedding layer. As the tweets vary in their lengths and raw text cannot be directly represented as dense vectors like images, we used padding and an extra “unknown” token during tokenization to achieve the fixed length input and represent any unseen tokens. As a potential avenue for follow up work, we encourage researchers to try different model architectures and pre-trained word embeddings to improve the model performance and learn new information.

## Results

### Data Records

The dataset is available at: https://figshare.com/s/7ce063c3d713adab8764 and is presented in two comma-separated value (csv) files, one collected from 17,323 autistic individuals with 3,137,952 tweets and the other collected from 171,273 control group users consisting of 3,377,518 tweets. Both the datasets have columns: User ID (a unique value assigned to each Twitter account), profile description (a short summary of the account posted by the user), account created (datetime when the account was created), friends count (number of accounts the user follows), followers count (number of account the user is being followed by), tweet date (datetime when the tweet was posted), tweet ID (a unique ID assigned to each tweet), language in which the tweet was posted, tweet text (original tweet), a list of hashtags present in each tweet, location (specified in the user profile, if the user provided one), number of replies (number of times the tweet has been replied to), number of retweets (number of times the tweet was retweeted), number of likes the tweet got, and source from where the tweet was posted (web, mobile device or app).

### Technical Validation

For tweet and user classification, the selection of an appropriate metric for evaluating the performance of a machine learning model is task-dependent and application-specific. Given that the present study pertains to a classification problem, we utilized accuracy, F1 score and AUC-ROC score as evaluation metrics. The results obtained from our tweet and user classifier is summarized below:

**Table 1.** Summary of results obtained for tweet classification from TF-IDF vectorization to identify best algorithm based on accuracy.

| Word vectorization method | Model | Validation set accuracy |
| --- | --- | --- |
| TF-IDF | SVM | 0.615 |
|  | Naive Bayes | 0.598 |
|  | Logistic Regression | 0.63 |
|  | XGBoost | 0.624 |

**Table 2.** Summary of results obtained for tweet classification from word2vec model using best identified algorithm.

| Word vectorization method | Model | Metric performance on test set |
| --- | --- | --- |
| Word2Vec | Logistic Regression | Accuracy: 0.73 |
|  |  | F1 score: 0.71 |
|  |  | AUC score: 0.728 |

While the TF-IDF vectorization yielded similar accuracy using different ML algorithms for tweet classification, logistic regression was chosen as the best predictor due to its superior performance and shorter training time. The results of the word2vec model were found to be consistent with the semantic similarities of the words. For instance, the word “autism” was found to have a higher cosine similarity to words such as “Aspergers”, “neuroatypical”, and “autism spectrum condition”. This suggests that the word2vec model was able to capture the semantic relationships between the words.

**Table 3.**
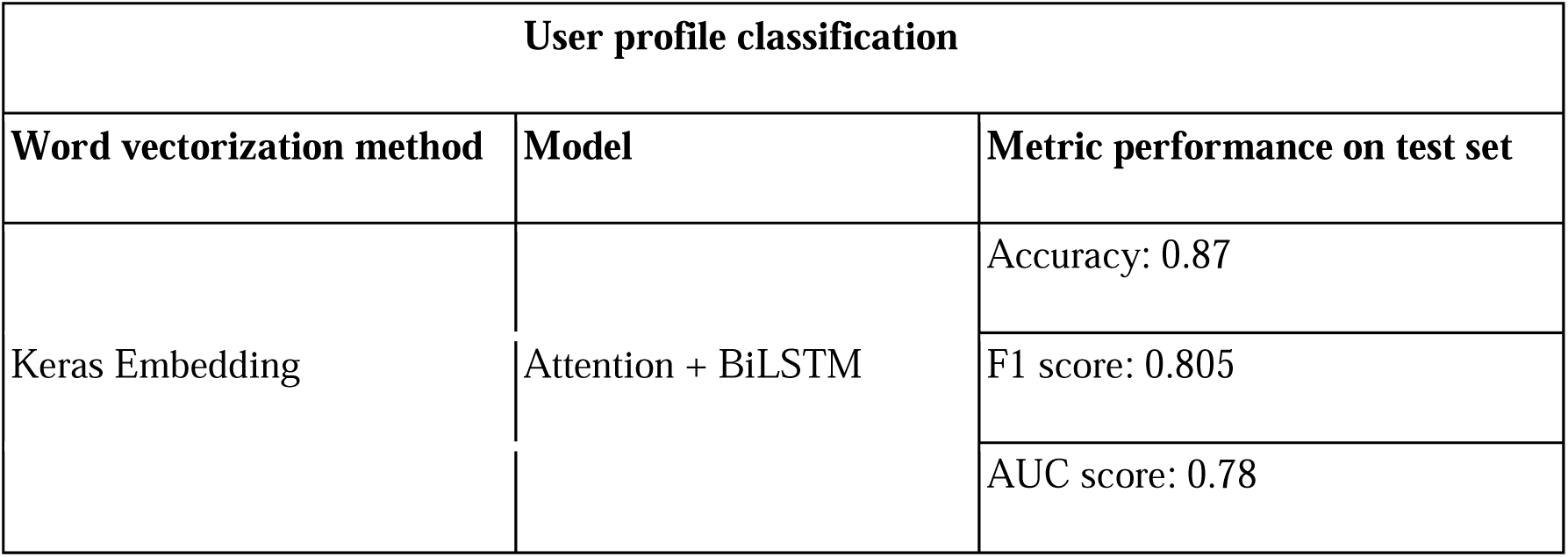
Summary of results obtained for user classification.

| User profile classification |  |  |
| --- | --- | --- |
| Word vectorization method | Model | Metric performance on test set |
| Keras Embedding | Attention + BiLSTM | Accuracy: 0.87 |
|  |  | F1 score: 0.805 |
|  |  | AUC score: 0.78 |

Although there is a class imbalance in the number of users found in ASD and control group dataset, the attention-based LSTM model still seems to make good predictions yielding an F1 score of 0.7 and 0.9 on the “autism” and “control group” classes respectively as well as an AUC score of 0.78. The results for topic modeling and sentiment analysis as obtained from the tweets using Top2Vec algorithm and VADER respectively are discussed in the Supplementary File 1.

## Discussions

We demonstrate the potential of using data mining techniques to learn about ASD and related topics from social media platforms such as Twitter and its ability to transform healthcare. The 73% accuracy achieved in the tweet classification and 87% in user classification shows that there are significant semantic differences in the messages posted by individuals with and without ASD. This finding, along with previous studies using computer vision models^61,66^ suggests that social phenotypical behavior could be used to support effective ASD screening strategies and facilitate early detection. Our dataset can facilitate valuable insights into various topics of discussion and the associated sentiments, carrying significant implications for public health decision-making, policy formulation, and clinical practice. The ability to observe behavioral symptoms in a non-clinical environment could lead to streamlined interactions between clinicians and patients, consequently enhancing both autism research and healthcare efficiency. We encourage researchers to benchmark the performances on this dataset and enhance the results using various techniques, or even combine our findings with research works similar to aforementioned works to build a multi-modal analytical tool. Such multimodal digital phenotyping methods have the potential to improve grading quality of clinical tools and shift healthcare from a reactive, disease-based model to a proactive, prevention-based model.

There are certain limitations to consider as well. While this study focused on individuals who self-identified as autistic, there is no clinical validation for their diagnosis. This is where annotations from clinical experts or crowdsourcing can help. Furthermore, there is a possibility of data leakage, where the identified users may not be autistic but instead could be family members, parents, caregivers, or advocacy organizations belonging to a different study population and still using the hashtags. Moreover, using VADER to label the emotional intensity and sentiments of the tweets can be relatively inaccurate compared to human labels, whose sentiments tend to get affected by their surroundings, politics and other factors, thus making it difficult to provide reliable labels. Additionally, this study only considered the English language, potentially missing out on information from other countries or languages that could aid the model in making better predictions. This also raises concerns of the lack of diversity in the data, where only English-speaking users from higher socio-economic groups or younger adults are represented in the dataset, as they comprise a larger portion of Twitter users.

This study presents several opportunities for future research, such ase using pre-trained large language models like BERT and GPT for text classification, topic modeling, and feature extraction. Another interesting avenue is the integration with additional data modalities such as audio and video which could also be mined from social media. In addition, incorporating auxiliary information to textual features may further improve the effectiveness of machine learning models. Lastly, as the Centers for Disease Control and Prevention has reported that boys are four times more likely to receive an ASD diagnosis than girls^67^, gender analysis using crowdsourcing or other metadata analysis techniques may also hold promise for future investigations.

## Data Availability

The curated dataset is available online at: https://figshare.com/s/7ce063c3d713adab8764 and https://www.kaggle.com/datasets/aditijaiswal/autism-twitter-dataset

https://figshare.com/s/7ce063c3d713adab8764

## Code Availability

We used standard Python packages such as Natural Language Toolkit (NLTK), matplotlib, and numpy for data preprocessing and analysis as well as tensorflow and scikit-learn for classification. The reference code used for tweet and user profile classification is available on GitHub (https://github.com/jaiswal-aditi/Twitter-Autism-Data.git)

## Acknowledgements

The technical support and advanced computing resources from University of Hawaii Information Technology Services – Cyberinfrastructure, funded in part by the National Science Foundation CC* awards # 2201428 and # 2232862, are gratefully acknowledged.

## Author Information

### Contributions

Aditi Jaiswal - data collection, data analysis, manuscript writing - original draft

Dr Peter Washington - conceptualization, supervision, manuscript reviewing and editing

