## Supplementary File 1 for "#ActuallyAutistic Twitter dataset for precision diagnosis of Autism Spectrum Disorder (ASD)"

**Exploratory Data Analysis**

We observed that the hashtags and location columns of the dataset contained the most number of missing values. While not all the tweets have a hashtag or cause associated with it, users have the freedom to enter any location they desire on their profiles. Our analysis revealed that the majority of users did not enter their actual location and those who did had inconsistent location entries. Of the top 20 location values found, the majority were variations of "United Kingdom", including "UK", "London, England", "England, United Kingdom", "South East, England", while others were less informative strings such as "Picnic party" and "My parent's basement".

Further analysis of the yearly distribution of tweets revealed an increase in ASD conversations over the years, as shown in supplementary figure S1. Autistic individuals appear to be more comfortable with social media interactions, which provides them with multiple employment opportunities and serves as an effective platform for educating people about developmental delays and sharing behavioral symptoms that may be helpful to others.

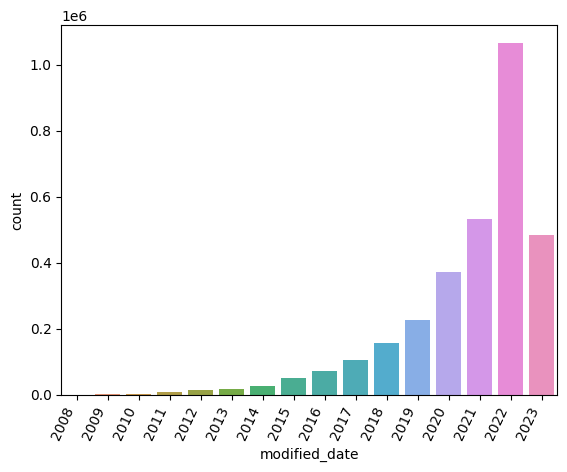

#### **Supplementary Figure S1.** Timely distribution of tweets posted by ASD users. Note that the count for 2023 is incomplete as it also collected additional data, from timelines of the users, up to and including February 2023.

**Sentiment Labeling**

Sentiment analysis has been extensively researched in NLP academic literature and is commonly studied using two approaches: machine learning and lexical. For this work, we utilized the Valence Aware Dictionary for sEntiment Reasoning (VADER)^1^, a lexical based approach *specifically attuned to sentiments expressed in social media or microblogs like context* to analyze the sentiments of the curated dataset. It has been explicitly attuned to social media datasets (like social media posts, New York Times editorials) and requires no training data. VADER applies a set of rules and heuristics on the sentiment scores of the individual words to determine the overall sentiment of the sentence and returns a dictionary of negative, neutral, positive, and overall (normalized) sentiment scores of the sentence. Our analysis focused on comparing the sentiments of tweets posted by individuals with ASD against those from the control group. However, an additional analysis was conducted on the original profane dataset as well to assess the potential impact of profanity on sentiment. But the released dataset only contains clean tweets with no profanity.

The VADER sentiments of most of the ASD and control group tweets were found to be positive and neutral respectively, as shown in supplementary figure S2.

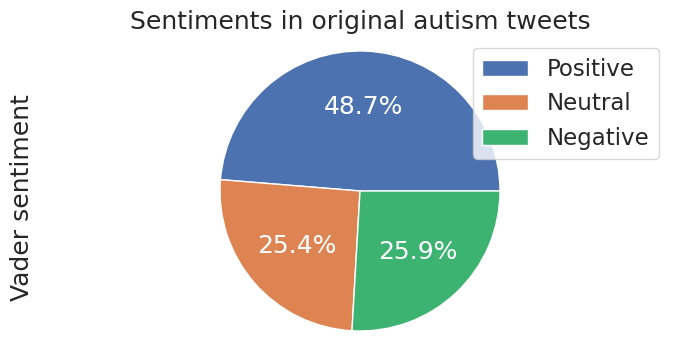

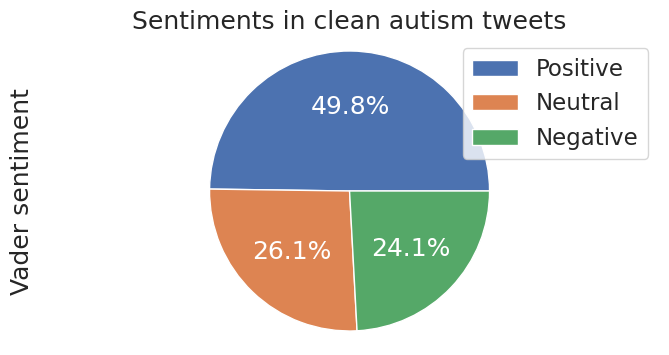

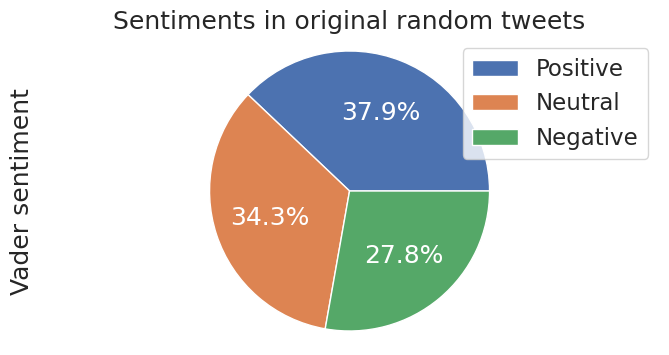

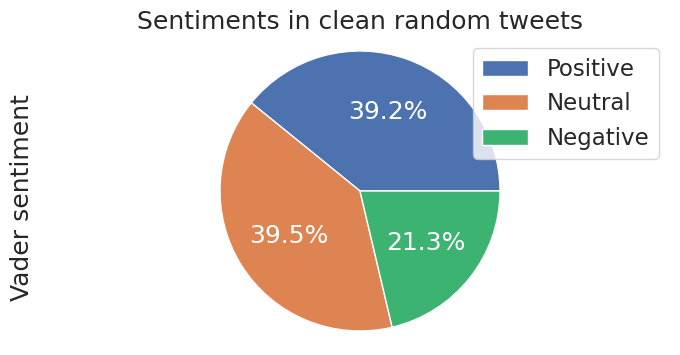

#### **Supplementary Figure S2.** Distribution of sentiments in the ASD and control group dataset

This was supported by another interesting observation, where the ASD tweets had more characters as compared to the control group tweets, shown in supplementary figure S3. The histograms of the tweet word counts of both the groups follow similar distributions, but with a substantial difference in their means. This is clearly indicative of the differences in linguistic patterns between the two groups.

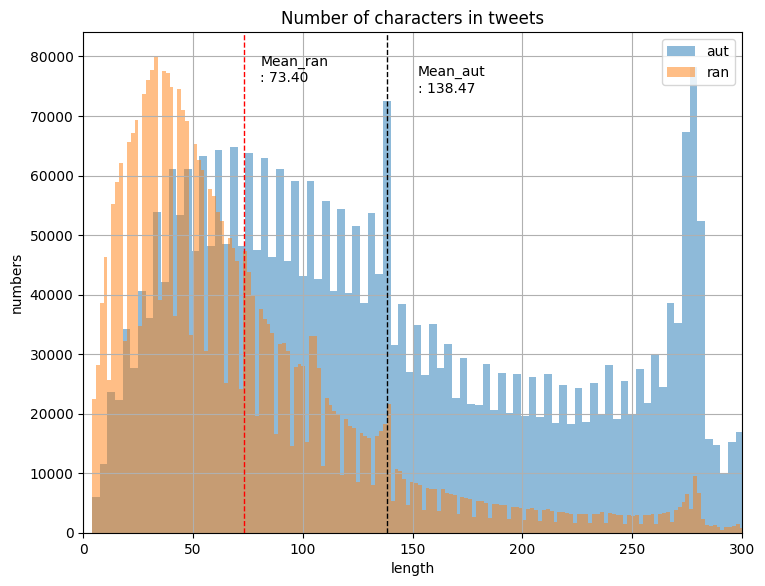

**Supplementary Figure S3.** Histograms of tweet character and word counts of the two groups

**Topic Modeling**

Topic modeling is an unsupervised learning technique used to identify hidden topics and structures in the documents and group related words representing coherent themes or topics. In our study, we employed the Top2Vec^2^ algorithm that offers a dynamic approach to discovering topics within a corpus of text data by making use of the spatial proximity of the words. Instead of a fixed number of topics, Top2Vec generates joint embedding of document and word vectors that capture the underlying semantic structure of the data. This allows similar documents and words to be grouped together, leading to more coherent and interpretable topics. To avoid a sparse vector representation, Top2Vec uses uniform manifold approximation and projection (UMAP) dimensional reduction, followed by a hierarchical density-based spatial clustering of applications with noise (HDBSCAN) clustering. This reserves the local and global structure of the vector space and finally the topic vectors are extracted from the centroids of the dense vectors.

The objective of our topic modeling analysis was to investigate whether there exist specific themes that are frequently discussed in relation to autism. Using just the ASD dataset, multiple topics were discovered and the word clouds of a few topics are shown in supplementary figure S4.

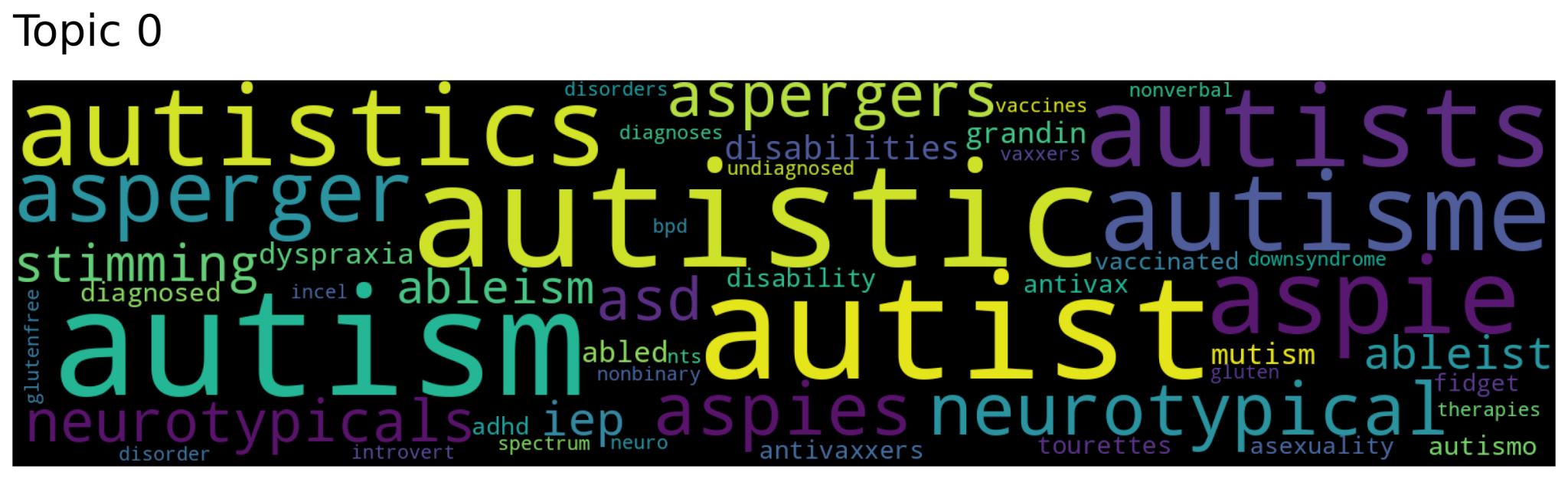

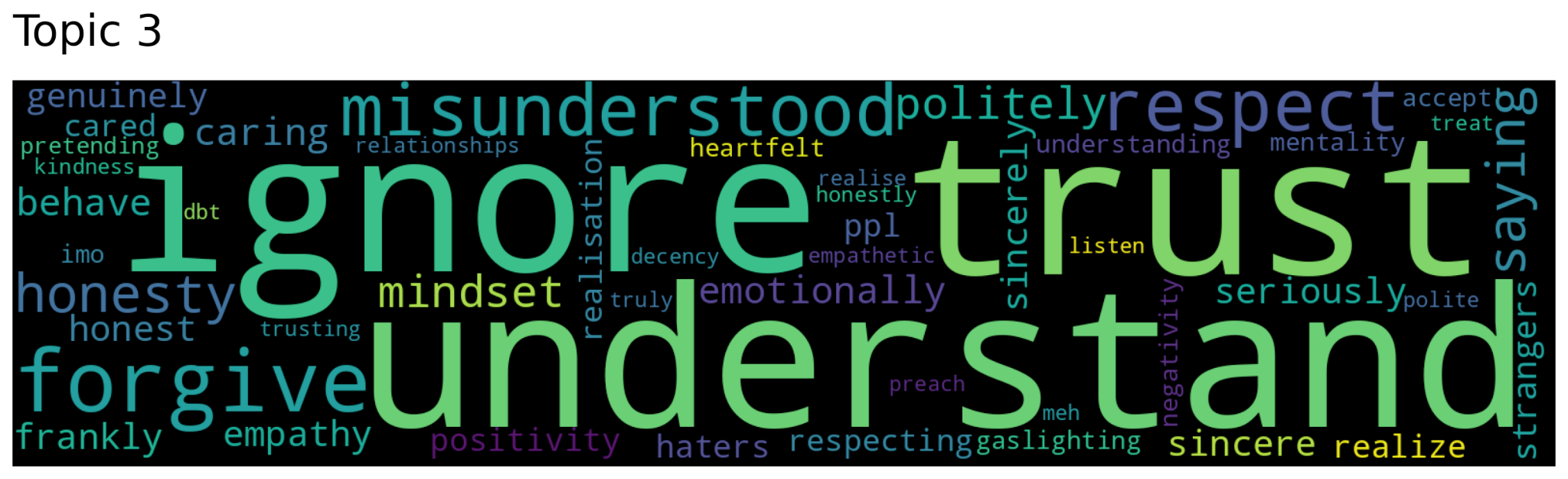

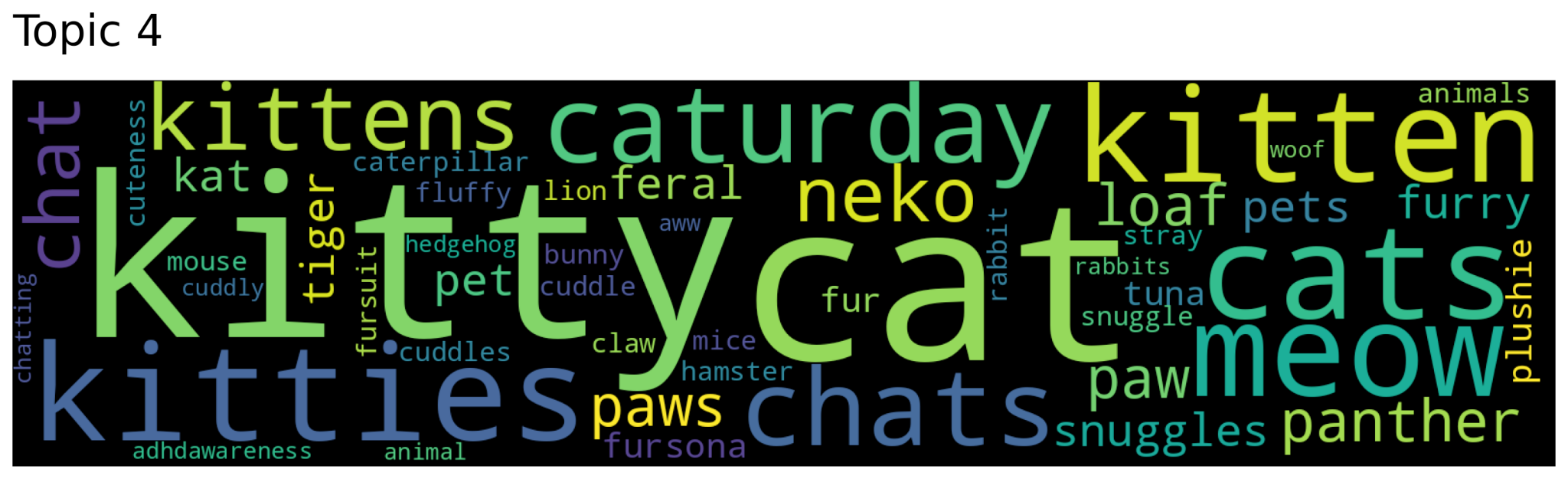

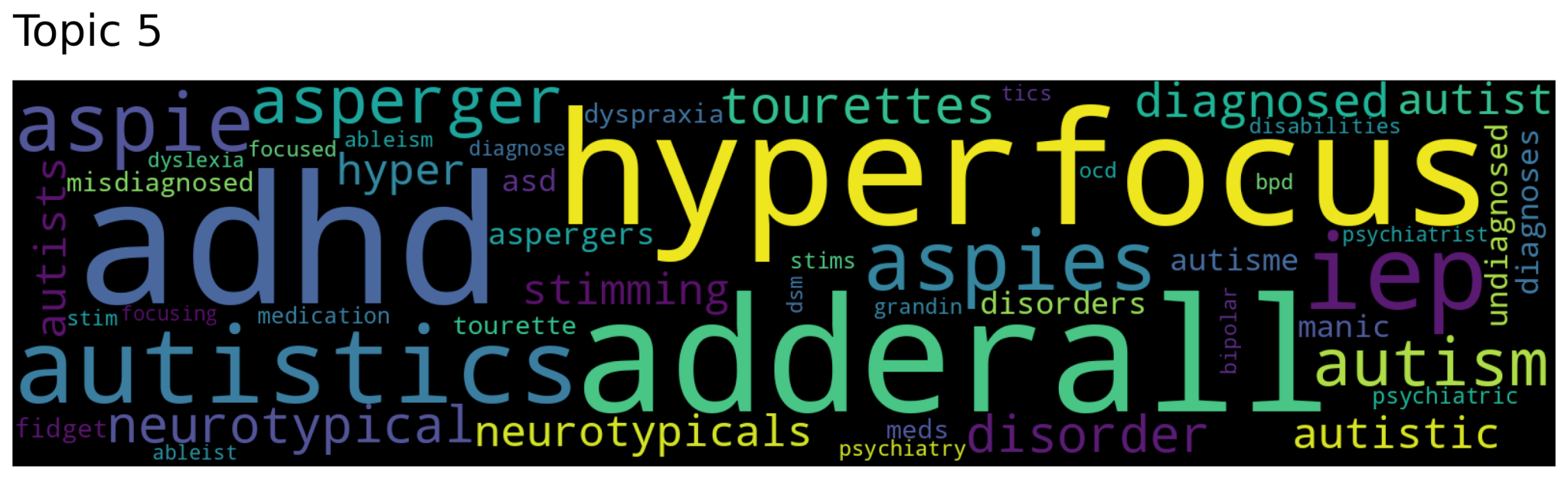

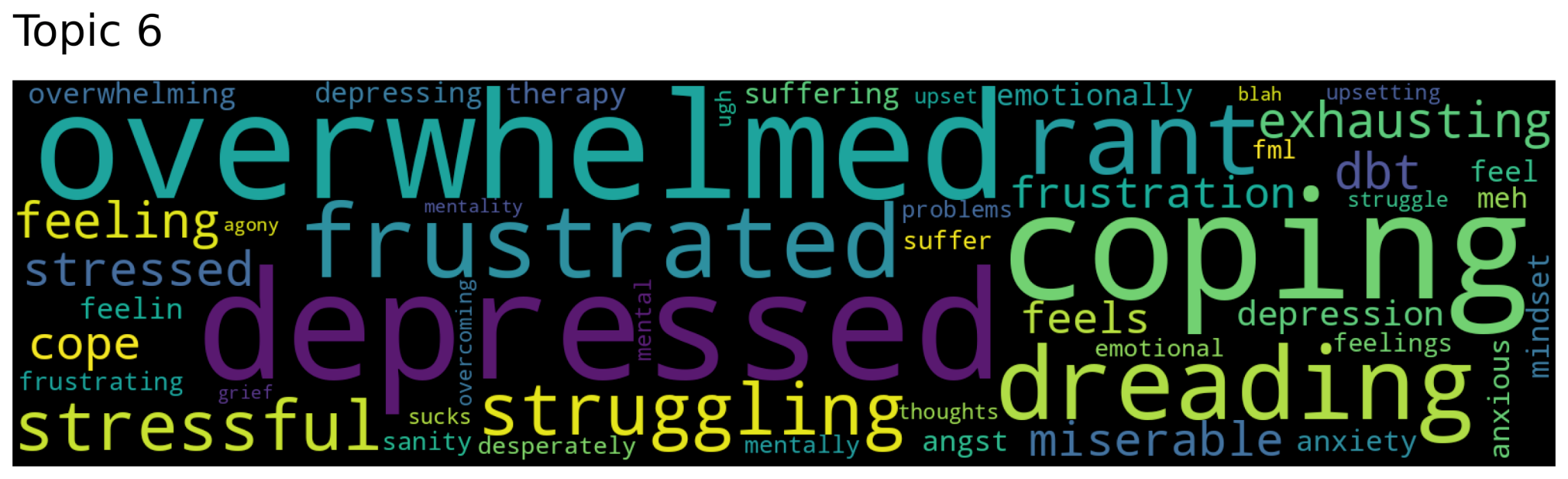

**Supplementary Figure S4.** Topics observed in ASD dataset using Top2Vec algorithm

The majority of topics were related to behavioral and emotional symptoms, such as "hyperactivity", "fidgeting", "depressed", "anxiety", "trembling", and "overwhelmed". Interestingly, a considerable number of documents were also focused on "vaccine", "therapy", "misdiagnosis", and "cats". These observations could be attributed to the frequent misdiagnosis or delayed diagnosis of autism, prompting individuals to seek therapy, support, and guidance. The presence of vaccine-related discourse is likely due to vaccine misinformation and its adverse impact on individuals affected by ASD. However, given the timeframe in which the dataset was collected, it is also possible that these tweets are related to COVID-19 vaccines. Lastly, multiple studies^3,4^ have found that autistic children are more at ease with cats, as they are less intrusive, do not maintain prolonged eye contact, and can help to relieve stress and understand emotional cues.

On the contrary, deriving specific topics from the control group's Twitter conversations was challenging, given their scattered and diverse nature. Most of these discussions centered around internet personalities, random conversations or specific days of the week or special occasions like birthdays and anniversaries. Surprisingly some topics were found to be related to animals, in general, as opposed to just cats which was observed in autistic user conversations. Some of these posts also displayed usage of emotional words suggesting that pets or animals may provide therapeutic benefits.
